## Supplementary Methods and Figures for "Aggregative *trans*-eQTL analysis detects trait-specific target gene sets in whole blood"

### Supplementary Text

#### Supplementary Section A. Methods

**A1. Extended methods.** To introduce the details of ARCHIE, first we briefly describe standard canonical correlation (CC) analysis. For *n* individuals, let $\boldsymbol{G}^{n\times p}$ be the normalized genotype matrix for *p* variants and $\boldsymbol{E}^{n\times g}$ be the normalized gene-expression matrix for *g* genes all of which are distant (*trans*) to the variants. The classical CC analysis seeks to find linear combinations of variants ($\boldsymbol{u}^{p\times 1}$) and genes ($\boldsymbol{v}^{g\times 1}$) such that the correlation between $\boldsymbol{Gu}$ and $\boldsymbol{Ev}$ is maximized i.e.,

$$(\boldsymbol{u},\boldsymbol{v}) = \boldsymbol{argmax}\, \widetilde{\boldsymbol{v}}^{\boldsymbol{T}}\boldsymbol{E}^{\boldsymbol{T}}\boldsymbol{G}\widetilde{\boldsymbol{u}}$$

$$\text{with } ||\widetilde{\boldsymbol{u}}||_{\boldsymbol{2}} = \boldsymbol{1} \text{ and } ||\widetilde{\boldsymbol{v}}||_{\boldsymbol{2}} = \boldsymbol{1}$$

where $||.||_{\boldsymbol{h}}$ denotes the $L_h$ norm. The subsequent pairs of CC components are obtained by similarly maximizing the correlation between $\boldsymbol{Gu}$ and $\boldsymbol{Ev}$ and under the constraint of being uncorrelated or orthogonal to the previous components. In practice, this can be obtained from the summary statistics of standard trans-eQTL mapping. If $(\boldsymbol{u_k},\boldsymbol{v_k})$ denote the $\boldsymbol{k^{th}}$ pair of CC components, then

$$\boldsymbol{u_k} = \boldsymbol{k^{th}} \text{ eigen-vector of } \boldsymbol{W^TW}$$

where $\boldsymbol{W} = \boldsymbol{\Sigma}_{\boldsymbol{EE}}^{-1/2}\boldsymbol{\Sigma}_{\boldsymbol{GE}}\boldsymbol{\Sigma}_{\boldsymbol{GG}}^{-1/2}$ and $\boldsymbol{v_k} = \boldsymbol{Wu}$ ; $\boldsymbol{\Sigma}_{\boldsymbol{GG}}$ and $\boldsymbol{\Sigma}_{\boldsymbol{EE}}$ are the column-correlations of $\boldsymbol{G}$ and $\boldsymbol{E}$ respectively and $\boldsymbol{\Sigma}_{\boldsymbol{GE}}$ is the cross-covariance matrix between the variants and the gene-expressions. In practice, we approximate $\boldsymbol{\Sigma}_{\boldsymbol{GG}}$ and $\boldsymbol{\Sigma}_{\boldsymbol{EE}}$ by the empirical LD-matrix and a penalized co-expression matrix (See next section) using external reference samples and $\boldsymbol{\Sigma}_{\boldsymbol{GE}}$ can be obtained from the summary statistics of the regression for trans-eQTL mapping across all pairs of variants and gene-expressions (See next section). Thus, although the objective function involves the individual data matrices $\boldsymbol{G}$ and $\boldsymbol{E}$, in practice the solution cab be obtained based on two sets of summary association statistics and external reference samples.

The CC components represent latent linear factors which explain the correlation between the variants and the gene-expressions by aggregating multiple, possibly weaker, associations. However, it is difficult to interpret the CC components directly since all variants and gene-expressions would have non-zero coefficients in the corresponding components for all pairs $(\boldsymbol{u_k}, \boldsymbol{v_k})$. For better interpretation, we proposed to derive CC components that will involve only limited number of SNPs and genes based on sparse canonical correlation analysis (sCCA). We introduce penalty terms to regularize the CC components thus effectively reducing the smaller components to 0 for a suitable penalty. In particular we use an $L_1$ penalty on each of the CC components, similar to LASSO. The optimization problem we solve is given by

$$(\boldsymbol{u}, \boldsymbol{v}) = \boldsymbol{argmax}\, \boldsymbol{\tilde{v}^T W \tilde{u}}$$

$$\text{with } ||\boldsymbol{\tilde{u}}||_{\boldsymbol{1}} \ \leq \ \boldsymbol{c_u}\ ;||\boldsymbol{\tilde{v}}||_{\boldsymbol{1}} \ \leq \ \boldsymbol{c_v} \text{ and } ||\boldsymbol{\tilde{u}}||_{\boldsymbol{2}} \ = \ \boldsymbol{1}, ||\boldsymbol{\tilde{v}}||_{\boldsymbol{2}} \ = \ \boldsymbol{1}$$

We have used $L_1$ penalty in this context as this is known to produce sparse solution and thus allows for variable selection. The resulting pair of sCC components $(\boldsymbol{u}, \boldsymbol{v})$ denote the coefficient loadings of the corresponding variants and genes respectively. A non-zero element in $\boldsymbol{u}$ (or $\boldsymbol{v}$) implies that the respective variant (or gene) was selected.

We solve the above optimization problem using an alternating maximization approach[1]. We note that for a fixed $\boldsymbol{u} = \boldsymbol{u_0}$, the objective function is similar to a standard LASSO as:

$$\boldsymbol{v} = \boldsymbol{argmax}\, \boldsymbol{\tilde{v}^T W u_0} \text{ with } ||\boldsymbol{\tilde{v}}||_{\boldsymbol{1}} \ \leq \ \boldsymbol{c_v} \text{ and } ||\boldsymbol{\tilde{v}}||_{\boldsymbol{2}} \ = \ \boldsymbol{1}$$

and similarly, for a fixed $\boldsymbol{u} = \boldsymbol{u_0}$

$$\boldsymbol{u} = \boldsymbol{argmax}\, \boldsymbol{v_0^T W \tilde{u}} \text{ with } ||\boldsymbol{\tilde{u}}||_{\boldsymbol{1}} \leq \boldsymbol{c_u} \text{ and } ||\boldsymbol{\tilde{u}}||_{\boldsymbol{2}} = \boldsymbol{1}$$

For given sparsity levels determined by sparsity parameters $\boldsymbol{c_u}$ and $\boldsymbol{c_v}$, the above optimization problems can be solved by alternating between $\boldsymbol{u}$ and $\boldsymbol{v}$ and using the soft-thresholding technique via the following algorithm

1) Initialize $(\boldsymbol{u}, \boldsymbol{v}) = (\boldsymbol{u_0}, \boldsymbol{v_0})$ with the first principal-components of $\boldsymbol{W^T W}$ and $\boldsymbol{W W^T}$ respectively.

2) Iterate until convergence:

   a) $\boldsymbol{v_t} \leftarrow \boldsymbol{argmax}\, \boldsymbol{v^T W u_{t-1}}$ with $||\boldsymbol{v}||_{\boldsymbol{1}} \leq \boldsymbol{c_v}$ and normalize $\boldsymbol{v}$ such that $||\boldsymbol{v}||_{\boldsymbol{2}} = \boldsymbol{1}$

   b) $\boldsymbol{u_t} \leftarrow \boldsymbol{argmax}\, \boldsymbol{v_{t-1}^T W u}$ with $||\boldsymbol{v}||_{\boldsymbol{1}} \leq \boldsymbol{c_u}$ and normalize $\boldsymbol{u}$ such that $||\boldsymbol{u}||_{\boldsymbol{2}} = \boldsymbol{1}$

3) cc-value is defined as $\boldsymbol{q^2} = \frac{(\boldsymbol{v^T W u})^{\boldsymbol{2}}}{\sqrt{(\boldsymbol{u^T W^T W u})(\boldsymbol{v^T W W^T v})}}$

Steps 2a and 2b can be solved using a soft thresholding operator $\boldsymbol{S}(\boldsymbol{a}, \boldsymbol{b}) = \boldsymbol{sign}(\boldsymbol{a})(|\boldsymbol{a}| - \boldsymbol{b})$ as described in Witten and Tibshirani[2]. Given the first pair of sparse CC components, we use the previous algorithm with a matrix deflated cross-covariance matrix to obtain the subsequent pairs of CC components

$$\boldsymbol{W_{deflated}} = \boldsymbol{W} - |\boldsymbol{q}|\boldsymbol{v u^T}$$

We note here that methods and applications based on sparse canonical correlations have previously been extensively studied in Witten et al. (2009) in context of penalized matrix decomposition. However, in its current form, ARCHIE has several differences with their approach: (1) ARCHIE corrects for LD and coexpression using $\mathbf{\Sigma}_{GG}$ and $\mathbf{\Sigma}_{EE}$ matrices respectively. In contrast, Witten and colleagues assumed these matrix to be diagonal. Thus, ARCHIE, by correcting for $\mathbf{\Sigma}_{GG}$ and $\mathbf{\Sigma}_{EE}$ can identify potentially independent trans-associations between the selected sets of variants and genes. (2) Although the software released by Witten et al can be applied to summary statistics, the formulation and estimation procedure is based on individual level data. In contrast, the proposed algorithm of ARCHIE uses only summary matrices obtained from publicly available sources.

**A2. Choice of sparsity parameters**

Most applications of variable selection use cross-validation techniques to determine the choice the tuning parameters that will maximize the prediction accuracy in an independent test sample. However, in our analysis, prediction is not the aim and interpretation of sCC components is of greater interest. Further, because of individual level data are typically not available, a cross-validation approach is not feasible. Below we propose choosing sparsity parameters based on an intersection-minimization approach to improve the interpretation of the sparse CC components.

Although traditional CC analysis produces orthogonal components, in sCCA orthogonality cannot be guaranteed by the estimation algorithm we used. Here, we aim to minimize the intersection of the variants (genes) selected in successive variant- (gene-) components to ensure that we capture approximately orthogonal patterns of *trans*-association. In particular, we use the following algorithm to estimate the tuning parameters:

Let, $\boldsymbol{c_{uk}}$ and $\boldsymbol{c_{vk}}$ , $\boldsymbol{k = 1, 2, 3, .., \min(p, q) - 1}$, are a set of grid points for the tuning parameters associated with variants and gene expression components, respectively.

1) For $\boldsymbol{k = 2, 3, .., \min(p, q)}$

a) Determine the largest $\boldsymbol{c_{uk}} \in [\mathbf{0}, \mathbf{1}]$ through a grid search such that $||\boldsymbol{u_k} \odot \boldsymbol{u_{k+1}}||_{\mathbf{0}}$ is minimized.

b) Determine the largest $\boldsymbol{c_{vk}} \in [\mathbf{0}, \mathbf{1}]$ through a grid search such that $||\boldsymbol{v_k} \odot \boldsymbol{v_{k+1}}||_{\mathbf{0}}$ is minimized.

2) $\boldsymbol{c_u} = \mathbf{max}(\boldsymbol{c_{uk}})$ and $\boldsymbol{c_v} = \mathbf{max}(\boldsymbol{c_{vk}})$

where $\odot$ denotes element wise vector multiplication. In steps 1a and b, $\boldsymbol{c_{uk}}$ and $\boldsymbol{c_{vk}}$ are determined using a two-way grid-search. We do not know of any theoretical results that might guarantee that the sets of variants (genes) can always be chosen to be mutually exclusive. However, in our analysis with the eQTLGen data, we obtained non-intersecting sets of selected genes and variants across the ARCHIE components making them orthogonal. Of note, in our sCCA framework, we have the additional flexibility of using different tuning parameters for different sparse CC components given by the $\boldsymbol{c_{uk}}$ and $\boldsymbol{c_{vk}}$ respectively.

**A3. Construction of the covariance matrices**

Estimating $\Sigma_{EE}$: To apply ARCHIE, we need to estimate $\Sigma_{EE}$ which denotes the covariance between the gene-expressions for the $g$ genes in our analysis. For this, we used individual level gene-expressions in whole blood for the GTEx (v8) participants. We first regressed the normalized gene-expressions of each of the $g$ genes on standard covariates like age, sex, genetic PCs and top 30 PEER factors and quantile normalized the residuals. However, since typically the number of genes ($g$) is much larger than the number of individuals, the standard covariance (or correlation) matrix estimate is rank deficient and hence not invertible. Hence, we estimated a $\Sigma_{EE}$ using a penalized covariance matrix[3] of the quantile-normalized residuals. Such shrinkage-based co-expression estimates have been previously used in estimate high-dimensional matrices in functional genomics applications and exploits the Ledoit-Wolf lemma[4,5] for analytic calculation of the optimal shrinkage intensity.

Estimating $\Sigma_{GG}$: We estimated $\Sigma_{GG}$ using sample linkage-disequilibrium ($r$) matrix constructed using the individual-level genotype data from 5,000 randomly chosen individuals of European descent in UK Biobank data[6]. However, if a large reference panel is not available, several techniques can be used to estimate $\Sigma_{GG}$ from publicly available datasets like 1000 Genomes.

1. SNPs on separate chromosomes can be considered independent and the LD for SNPs on each chromosome can be estimated separately. In eQTLGen datasets, usually this strategy results in the number of samples being much larger than the number of SNPs on each chromosome. This guarantees that the $\Sigma_{GG}$ matrix remains positive semidefinite.

2. If the number of SNPs on each chromosome exceed the sample size of the reference data, sparse LD estimation with L1 penalty can be used. This will result in a sparse LD matrix which retains the stronger LD values, while reducing the weaker ones to zero. There are several available methods to estimate sparse LD[3,7].

Estimating $\Sigma_{GE}$: $\Sigma_{GE}$ represents the cross-covariance matrix between the genetic variants and distal gene expressions. Given the summary statistics (Z-value, p-value) of the trans-eQTL mapping, we can estimate $\Sigma_{GE}$ as:

$$(\Sigma_{GE})_{\boldsymbol{ij}} \approx \frac{\boldsymbol{Z_{ij}}}{\sqrt{\boldsymbol{2Nm_i(1-m_i)}}}$$

where $\boldsymbol{Z_{ij}}$ is the Z-value for the trans-eQTL mapping (linear regression) of the i[th] gene expression and j[th] genetic variant, $\boldsymbol{m_i}$ is the minor allele frequency of the variant and N is the effective sample size. This relationship holds under the assumption that the variance of gene expression explained by the SNP is negligible. In our analysis, $\boldsymbol{Z_{ij}}$ and N are provided by the eQTLGen trans-eQTL summary statistics, and $\boldsymbol{m_i}$ is estimated from an external reference panel like UK Biobank or 1000 Genomes.

#### Supplementary Section B. Numerical experiments.

We performed a small-scale resampling experiment to evaluate the performance of ARCHIE in capturing downstream trait-specific effects.

##### B1. Identification of downstream genes.

To demonstrate that ARCHIE can identify downstream trans-associations, we simulate individual level gene-expression data for N individuals (N=1,000 or 30,000). The sample sizes were chosen to reflect the approximate effective sample sizes of GTEx v8 and eQTLGen studies respectively. First, we randomly sampled 50 SNPs from the UK Biobank data with MAF varying between 10-40%, at least 50 Kb apart, such that the LD (r2) between any pair of SNPs is less than 0.05. Then we simulated gene-expression data using a given causal model (Figure 2B, C or Supplementary Figure 3). We grouped successive SNPs to have a direct regulatory effect on a cis-gene expression (marked in red throughout). For example, SNPs 1-5 had direct regulatory effect (proxy “cis” SNPs) on the expression levels of one gene, SNPs 6-10 on that of another gene etc. The cis-gene expressions for 8 genes (marked in red in Fig 2A-C) were simulated from the following gaussian error model:

$$\boldsymbol{E_j} \sim \boldsymbol{N}(\sum_{i=1}^{k} \boldsymbol{\beta_i G_{ij}}, \boldsymbol{1})$$

Where $\boldsymbol{E_j}$is the expression level of a cis gene in the $j^{th}$ individual, $\boldsymbol{G_{ij}}$ is the standardized genotype at the $i^{th}$ SNP of the $j^{th}$ individual and $\boldsymbol{\beta_i}$ is the corresponding direct effect of the $i^{th}$ SNP. We set

$\boldsymbol{\beta_i} \sim \boldsymbol{N(0.1, 0.04)}$ through for each SNP. We expect that for each cis-gene, the corresponding 5 regulatory SNPs explain a cis-heritability of approximately 20 - 22%.

For type-1 error simulations (Figure 2A), the downstream genes 1-9 were simulated independent of the cis genes. We simulated the expression levels of the downstream genes: Gene 1-9 using the cis gene expressions levels in a gaussian error model as follows:

$$\boldsymbol{Y_j \sim N(\sum_{i=1}^{k} \gamma_i X_{in;ij}, 1)}$$

Where $\boldsymbol{Y_j}$ is the expression of a downstream gene (Gene 1-9) for the j$^{th}$ individual, $\boldsymbol{X_{in;ij}}$is the gene expression in j$^{th}$ individual of the i$^{th}$ gene which has direct causal effect on the downstream gene expression and $\boldsymbol{\gamma_i}$ is the corresponding direct effect. For example, in Figure 2B, the expression level of Gene 1 is regulated by two cis genes mediating the effects of SNPs 1-10; the expression level of Gene 6 is regulated by Gene 2 and Gene 3. For causal effects $\boldsymbol{\gamma_i}$ between genes, we sample $\boldsymbol{\gamma_i \sim N(0.1, v/r)}$ where $\boldsymbol{r}$ is the number of direct causal effect on the specific gene and $\boldsymbol{v}$ is the variance parameter that controls the heritability of gene expression explained. we choose $\boldsymbol{v}$ and $\boldsymbol{r}$ such that the total heritability of downstream Genes 1-9 explained by the SNPs 1-40 is maintained at 10-14%.

The principal aim of ARCHIE is to accurately identify Genes 1-9 which has indirect associations with multiple SNPs, but the effect of the SNPs might be substantially attenuated due to the intermediate gene network. In context of our application with eQTLGen data, ideally the SNPs 1-40 are associated with a particular trait or disease (curated from external GWAS). Thus Gene 1-9 would reflect genes which cumulates independent effects of multiple SNPs associated with the trait.

For applying ARCHIE, we first performed standard trans-eQTL mapping to generate the summary statistics. Further, since there is no LD between SNPs, $\boldsymbol{\Sigma}_{GG}$ was assumed to be a diagonal matrix while we generated an additional $\mathbf{N}_{Ref}$ samples to estimate the $\boldsymbol{\Sigma}_{EE}$ separately. Using this simulated data, we applied ARCHIE and compared the results to that obtained from standard trans-eQTL mapping for two different sample size (1,000 and 30,000).

**B2. Presence of master regulator**: In addition to the simulation scenarios presented in Figure 2, we further compared the power of ARCHIE with standard trans-eQTL mapping in presence of a master regulator gene in the causal regulatory network (Supplementary Figure 3). Here, we simulate gene expressions of 11 genes, under similar parameter choices as above (See previous section). Gene 6 is the master regulator gene in the network. We find that across different choices of the sample size, ARCHIE has similar or higher power to detect downstream causal genes (Genes 5-11) compared to standard trans-eQTL. The empirical power estimates were calculated from 10,000 iterations while calibrating for type-I error.

**B3. Specificity and Sensitivity of the selected genes**: We further performed a simulation to assess the sensitivity and specificity of ARCHIE in comparison to standard trans-eQTL mapping. We simulated a causal regulatory network similar to that presented in Figure 2B (sparse causal network). However, we now designate a subset of SNPs as identified SNPs of interest. Thus, the aim is to identify the genes within the network which are regulated by these SNPs of interest (marked in green). The difference with the scenarios presented in context of power estimation is that, in that case, all the SNPs 1-40 were of interest and hence each gene in the causal network was regulated by these SNPs. However, in the current scenario only a subset of genes in the network are regulated by the SNPs which we call the non-null genes (marked in green), and the rest are called the null genes (marked in blue). We apply ARCHIE with the SNPs of interest and the genes in the network to assess whether ARCHIE can identify the null and the non-null genes correctly. In particular, we define sensitivity by the proportion of simulation iterations where ARCHIE selects at least one non-null gene correctly and specificity by the proportion of simulation iterations where ARCHIE does not select all the null genes. The metrics are defined similarly for standard trans-eQTL mapping as well. Throughout we use levels such that the type-I errors of the ARCHIE and standard trans-eQTL remains similar.

The results (Supplementary Figure 3) show that for both the scenarios considered with 10 and 20 SNPs of interest and 3 and 6 non-null genes within the network of 9 genes, the sensitivity of ARCHIE and standard trans-eQTL remain comparable. This indicates that at similar type-I error rates ARCHIE and standard trans-eQTL can identify the non-null genes with similar probability. However, the specificity of ARCHIE is substantially higher than that of standard trans-eQTL. This means that the chances of false discovery within the gene network is substantially lower for ARCHIE as compared to the standard trans-eQTL mapping

**B4. Assessing trait-specificity.**

Next, we demonstrate through resampling experiments that ARCHIE can potentially identify trait specific trans associations. Using data from eQTLGen consortium, we construct a matrix ($A^{p\times g}$) of trans-eQTL summary statistics (Z-values) across for $p$ variants and $g$ genes. Out of the $p$ variants, we set $\delta$ proportion of them to be related to a particular trait. Thus, for high values of $\delta$ we expect that the matrix **A** would reflect trans-association patterns pertaining to the trait and hence should be captured by the ARCHIE components. For low values of $\delta$, the matrix **A** would reflect a competitive null situation, with the trans-association not pertaining to any particular trait. Using this matrix, **A**, along with the corresponding estimates of LD and co-expression as described above, we estimate the ARCHIE components and evaluate their significance. In particular, the quantity of interest is the empirical probability of at least one ARCHIE component to significant. To estimate that, we perform the above experiment multiple times for fixed values of p, g and $\delta$ and calculate the proportion of times at least one ARCHIE component is significant. We would expect ARCHIE to have higher probability of identifying significant components with increasing value of.

In our numerical experiments, we set p = 100, g = 5,000 throughout all the settings. We studied the empirical probability of at least one ARCHIE component to be significant with varying values of $\delta$. Given a particular trait, e.g., Type-1 Diabetes, for each value of $\delta$, we set $\delta$ proportion of the 100 (=p) variants to be related to Type-1 Diabetes. We further repeated the experiment with Height, Rheumatoid Arthritis and LDL Cholesterol (Supplementary Figure 6). For $\delta$=0, the estimated probability of at least one ARCHIE component to be significant is lesser than 0.0001. This indicates that the testing procedure against the competitive null hypothesis we adopted via the resampling algorithm outlined, can maintain type-I error. Further across the 4 different traits, we note that with increasing $\delta$, the estimated probability of at least one ARCHIE component to be significant increases and is greater than 80% when $\delta > 0.75$ for all the four traits. This indicates that with increasing trait-specificity in the matrix of summary statistics, ARCHIE has an increased probability of identifying a significant component by aggregating the corresponding weaker trans-associations.

B5. Variations due to reference sample size: We further investigated the impact of variation in $\mathbf{N}_{\boldsymbol{Ref}}$ (sample size of the transcriptomic data to estimate $\boldsymbol{\Sigma}_{\boldsymbol{EE}}$) on the power of ARCHIE. Using the causal model in Figure 2A and genotypes of 50 SNPs, we simulated gene expressions of 1,000 individuals. A separate dataset of $\mathbf{N}_{Ref}$ individuals were simulated to estimate $\mathbf{\Sigma}_{EE}$ to be used in the estimation of components in ARCHIE. Under this simulation setting, we varied from 100 to 700 to study how of the estimation of $\mathbf{\Sigma}_{EE}$ from the reference transcriptomic data impacts inference from ARCHIE. The results show that the highest power for ARCHIE is obtained approximately at 450 sample size and remain roughly constant thereafter. In general, robust performance can be achieved under an optimal ratio of the sample sizes of the reference transcriptomic study and the study from which summary statistics of standard trans-eQTL associations are being analyzed[8].

#### Supplementary Section C. Follow-up analysis.

**C1. Enrichment analysis**. To see if the selected genes in the gene-component of ARCHIE were enriched in pre-defined pathways, we used gene-set (pathway) enrichment analysis. In particular we applied hypergeometric (Fisher's exact) test to see if the selected genes are overrepresented in the pathways compared to what is expected at random. Further, to adjust for multiple testing, we use a false discovery rate (FDR) adjusted p-value for evaluating significance of the pathways. Gene-sets were obtained from numerous known and reported databases including Molecular signatures database[9,10] (MSigDB containing several known databases like KEGG, REACTOME, GO and Wiki Pathways), TRRUST[11], ITFP[12] and RegNetwork[13]. We used several commonly used online tools like FUMA and Shiny GO (See URL) for the analysis. Further, we obtained pre-computed lists of genes that are differentially expressed in a particular tissue for GTEx (v8) participants. Using a similar enrichment test, we investigated whether the gene-set selected by ARCHIE is enriched among these differentially expressed genes in each of the 54 tissues. Similar to gene-set (pathway) enrichment, we used also tested for interaction enrichment among the protein corresponding to the selected genes via hypergeometric test using the data from STRING (v11.0)[14].

**C2. Differentially expressed genes**. For each tissue, we curated lists of differentially expressed genes across the genome. To do this we used individual level gene expression data from GTEx v8 whole blood. We first adjusted each gene-expression using standard covariates like age, sex, genetic PCs and top 30 PEER factors and quantile normalized the residuals. Following this we performed a two-sided t-test for the expression of a gene in a particular tissue against the rest and this was done across all the genes. We defined a gene to be differentially expressed in a tissue, if the corresponding tissue-specific gene-expression was significantly different from that in the rest of the tissues, i.e., FDR adjusted p-value of the two-sided t-test was < 0.05. This resulted in a list of genes for each tissue representing the significantly differentially expressed set of genes in that tissue. The selected target genes were tested for enrichment against these lists of differentially expressed genes for each tissue using a hypergeometric test. All the genes used in each analysis was used as the corresponding background set for enrichment, i.e., for SCZ the analysis was restricted to a background set of 7,047 genes. If significant for a tissue, this would indicate that the selected set of target genes are overrepresented among the set of highly differentially expressed genes in the tissue, highlighting the possible role of the that tissue in the genetic etiology of the trait.

C3. Robustness of estimated $\boldsymbol{\Sigma}_{EE}$: To investigate the robustness of the results from ARCHIE, we performed an example analysis with Schizophrenia (SCZ). In the main analysis, we estimated the $\boldsymbol{\Sigma}_{EE}$ matrix from GTEx v8 Whole Blood. In this analysis, we replaced the $\boldsymbol{\Sigma}_{EE}$ estimated from gene expressions reported in Depressions genes and networks (DGN) study. This study in Battle et al. reports individual level gene expression levels in Whole Blood for more than 16,000 genes in 922 individuals. Similar preprocessing steps were performed with the gene expression data as outlined in Supplementary Section A3. Of the initial 7,047 genes considered in the main analysis with GTEx v8 (See Results), DGN reported expression levels of 6,919 genes. ARCHIE identified 72 genes through the gene component, out of which 62 (83%) were also identified in the main analysis with GTEx v8. Further, among the 59 novel genes identified in the main analysis, 45 (76%) were also selected in this analysis by ARCHIE. This demonstrates that the results of ARCHIE can be a robust to the choice of reference studies.

### Supplementary Figures

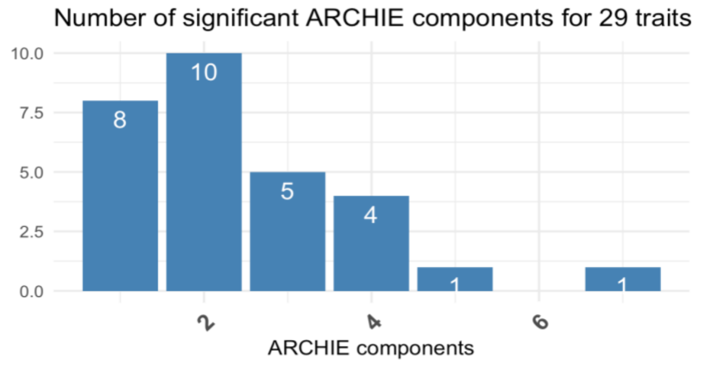


**Supplementary Figure 1**: Number of significant components identified by ARCHIE for variants associated 29 traits reported in eQTLGen consortium data.

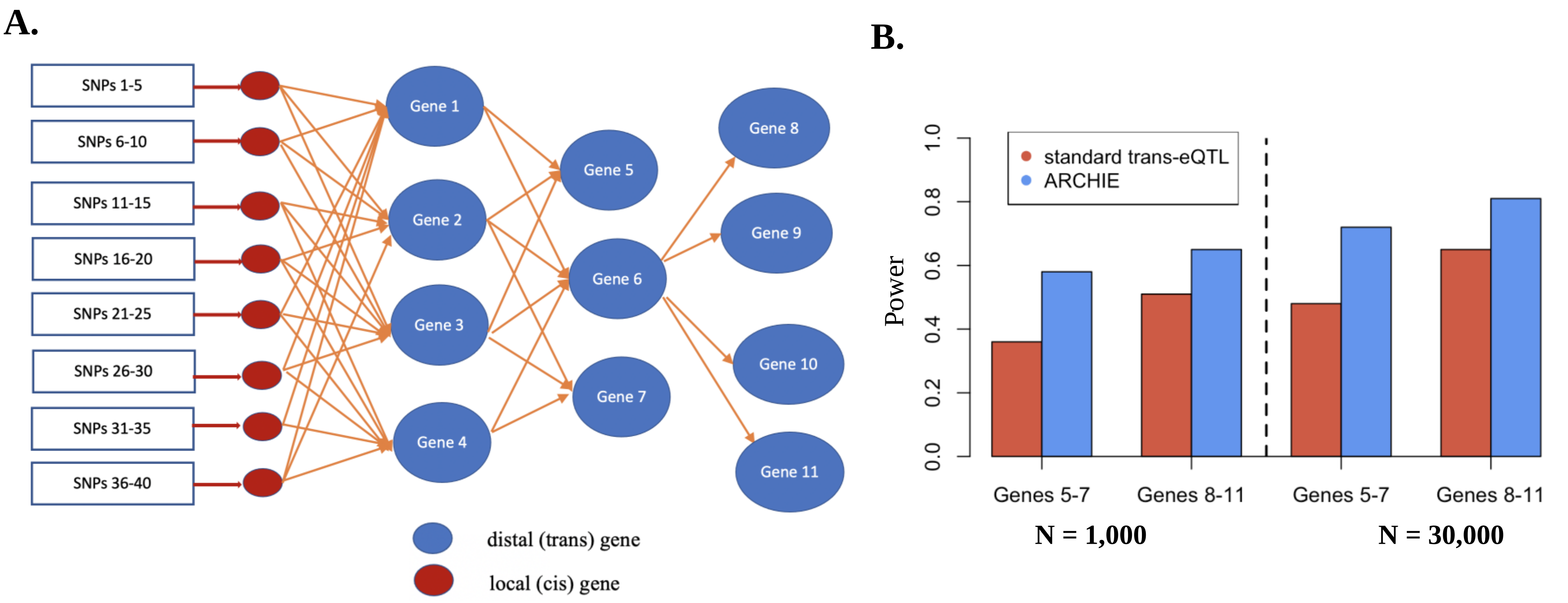


**Supplementary Figure 2. Power of ARCHIE in presence of master regulator. (A)** Causal regulatory network for associations between SNPs 1-40 and Genes 1-11. The red circles denote local (cis) genes, and the blue circles denote the distal (trans) genes. Orange arrows denote the causal effect of genes, and the red arrows denote the causal effect of SNPs on cis-genes. See Methods for more details on simulation model. Gene 6 is the potential master regulator in the causal network. **(B)** Empirical power estimates for standard trans-eQTL and ARCHIE to identify Genes 5-7 and Genes 8-11, calibrated to same type-I error level, across different sample sizes for the transcriptomic study.

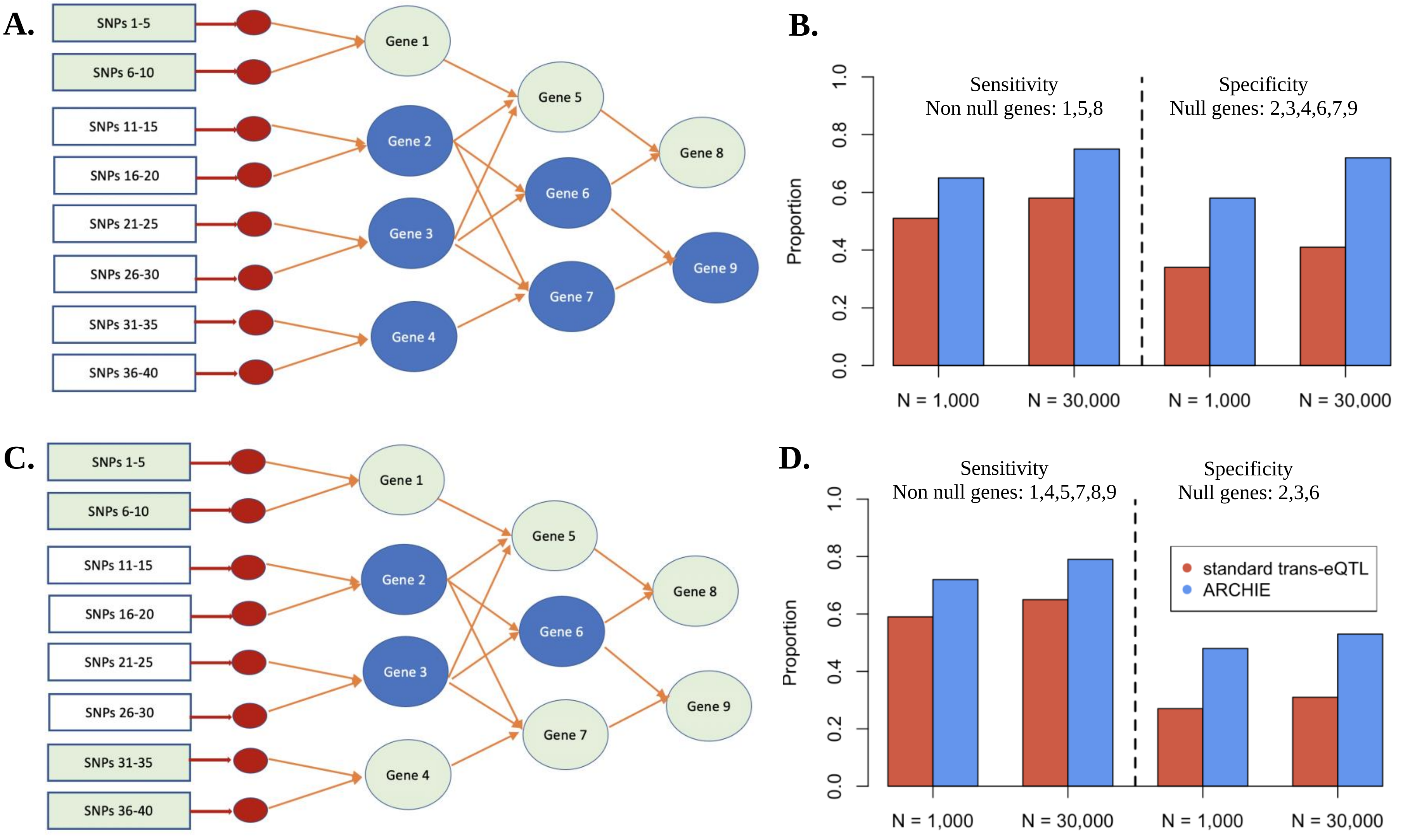


**Supplementary Figure 3. Sensitivity and Specificity of ARCHIE.** (A) Sparse causal regulatory network same as Figure 2B. We assume that SNPs 1-10 are observed/identified only, which means Genes 1, 5 and 8 are only causally regulated (non-null) and the rest of the are not (null genes). See Methods and Supplementary Methods for more details on simulation. (B) Estimated sensitivity and specificity of ARCHIE and standard trans-eQTL across two different sample sizes, calibrated for same type-I error. (C) Sparse causal regulatory network where SNPs 1-10 and 31-40 are observed only, which means Genes 1, 4, 5, 7 and 8 are causally regulated (non-null) and the rest of the are not (null genes). (D) Estimated sensitivity and specificity of ARCHIE and standard trans-eQTL across two different sample sizes, calibrated for same type-I error.

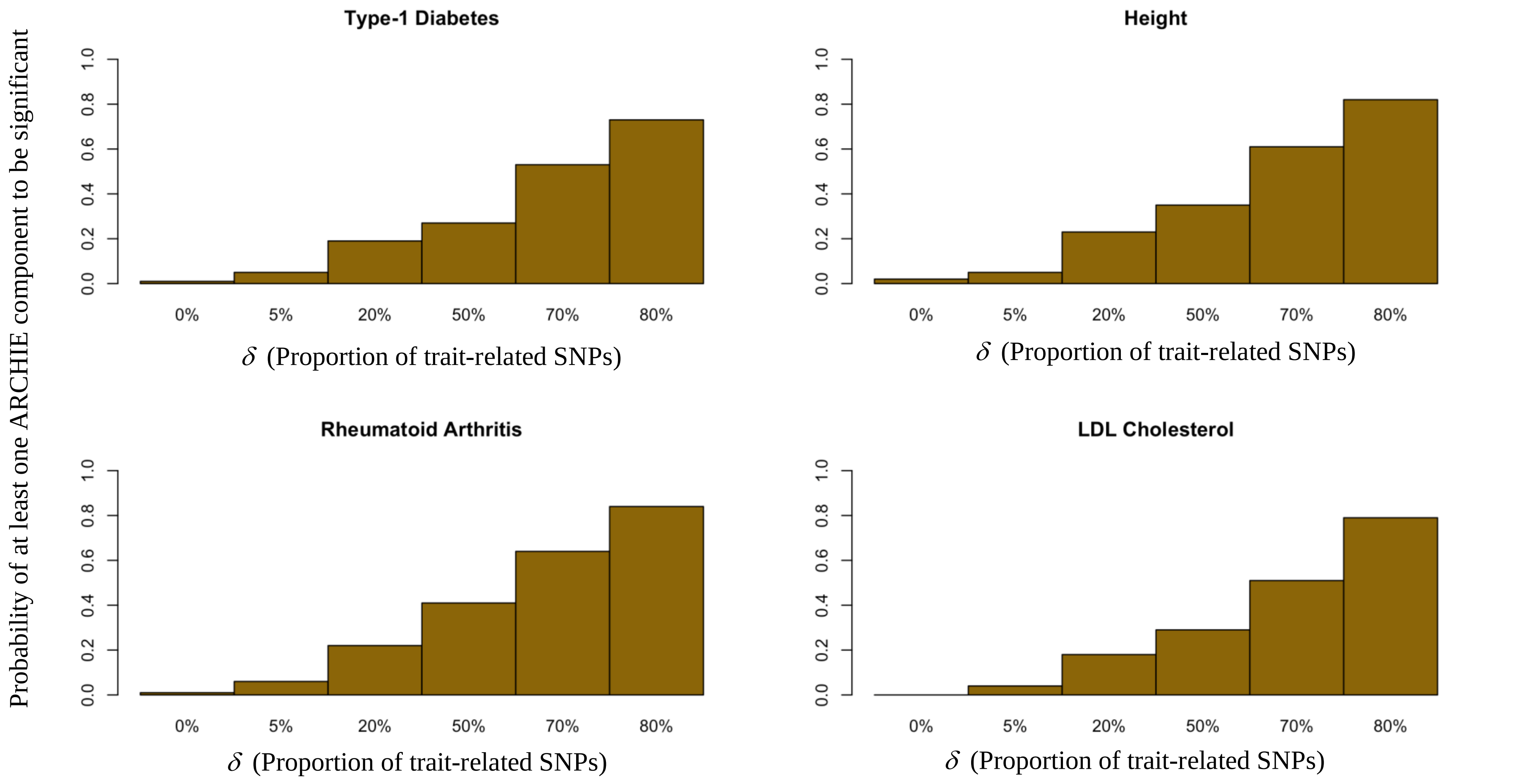


**Supplementary Figure 4. Type-I error and Power under competitive null hypothesis.** Probability of at least one ARCHIE component to be significant for 4 different traits in yellow bars across different values of $\delta$ (See Supplementary Methods for details on the resampling experiment). The significance level was set to at 0.0001

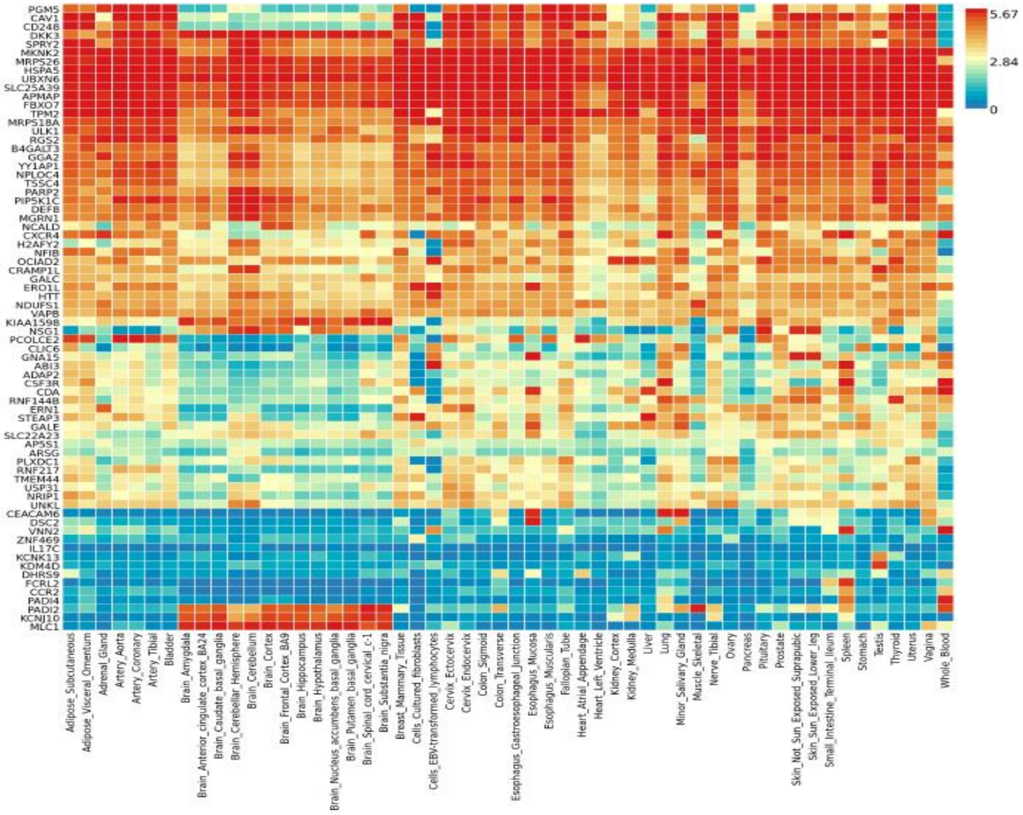


**Supplementary Figure 5.** Log2-transformed mean expression levels for 75 target genes selected by ARCHIE for SCZ across 54 GTEx v8 tissues. Three genes (*PADI2*, *KCNJ10*, *MLC1*; bottom 3 rows) are highly upregulated and three genes (*PGM5*, *CAV1* and *CD248*; top 3 rows) are highly downregulated particularly in brain tissues.

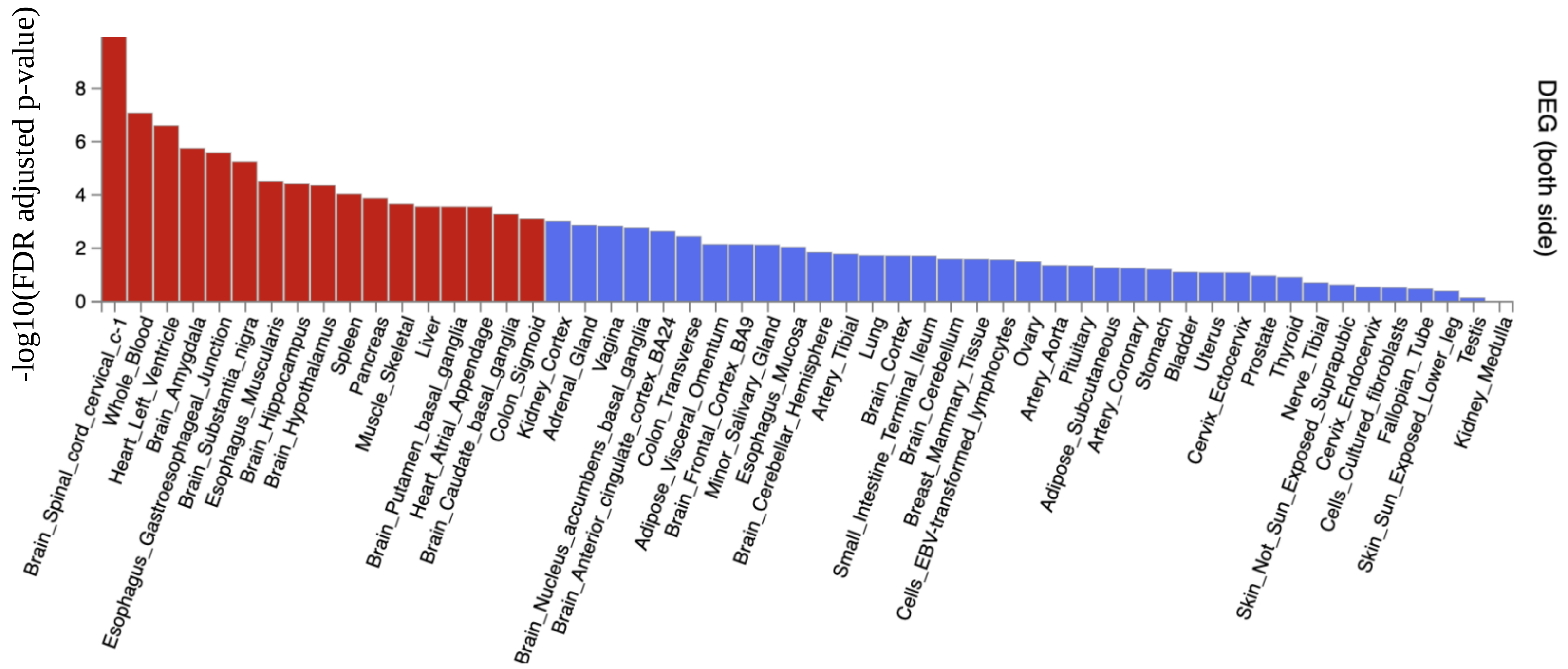


**Supplementary Figure 6.** Enrichment analysis with the 75 selected target genes for SCZ for enrichment among pre-determined list of differentially expressed genes (DEG; positive and negative) in each tissue of GTEx v8 and their corresponding enrichment p-value. The red bars denote the p-values for the tissues which are significant at FDR-adjusted p-value threshold of 0.001.

A.

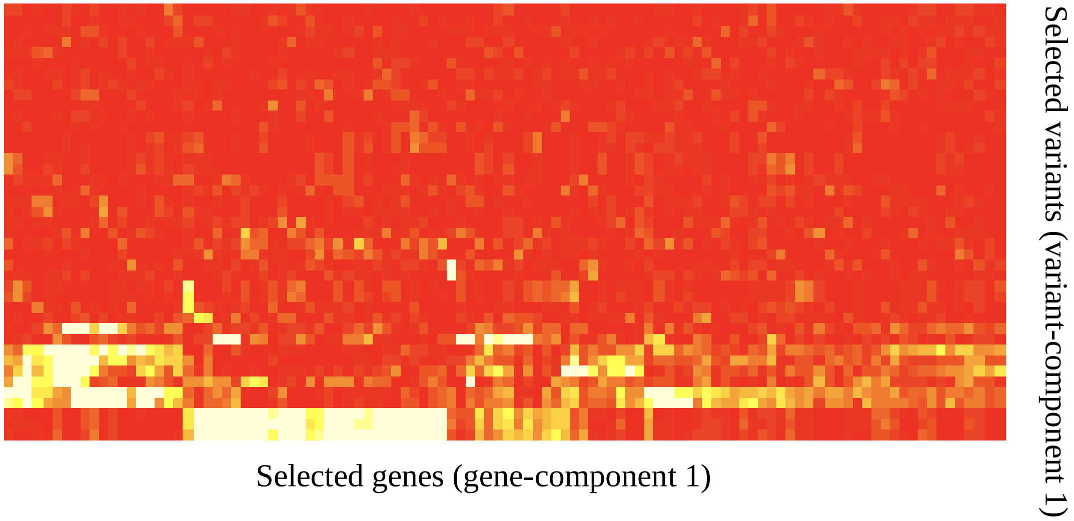


B.

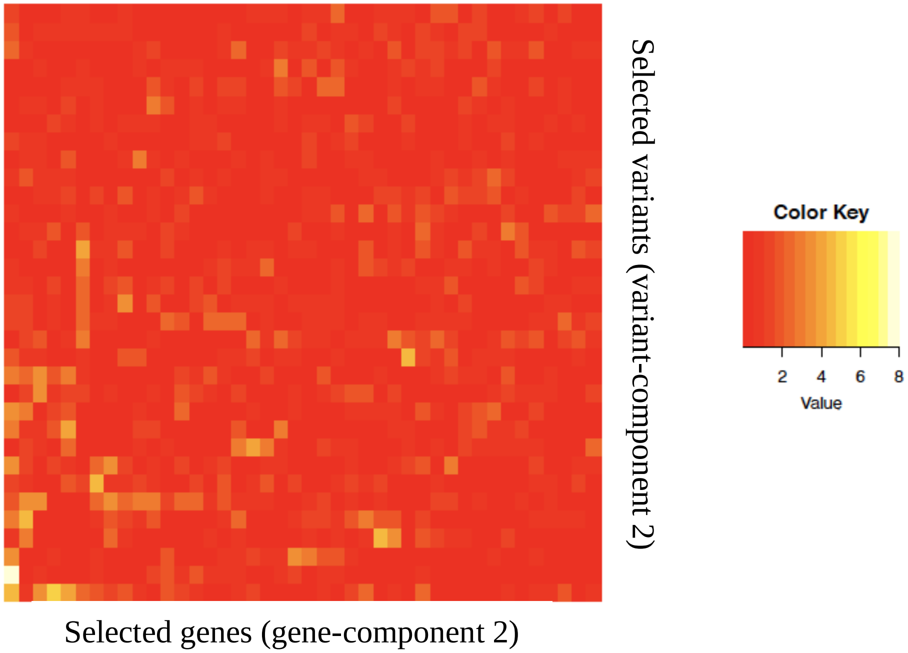


**Supplementary Figure 7.** -$\log_{10}$(p-values) for trans-eQTL (as reported in eQTLGen summary statistics) association between variants and genes selected in (A) ARCHIE component 1 and (B) component 2. Any association p-value < $10^{-08}$ is collapsed to $10^{-08}$ for the ease of viewing.

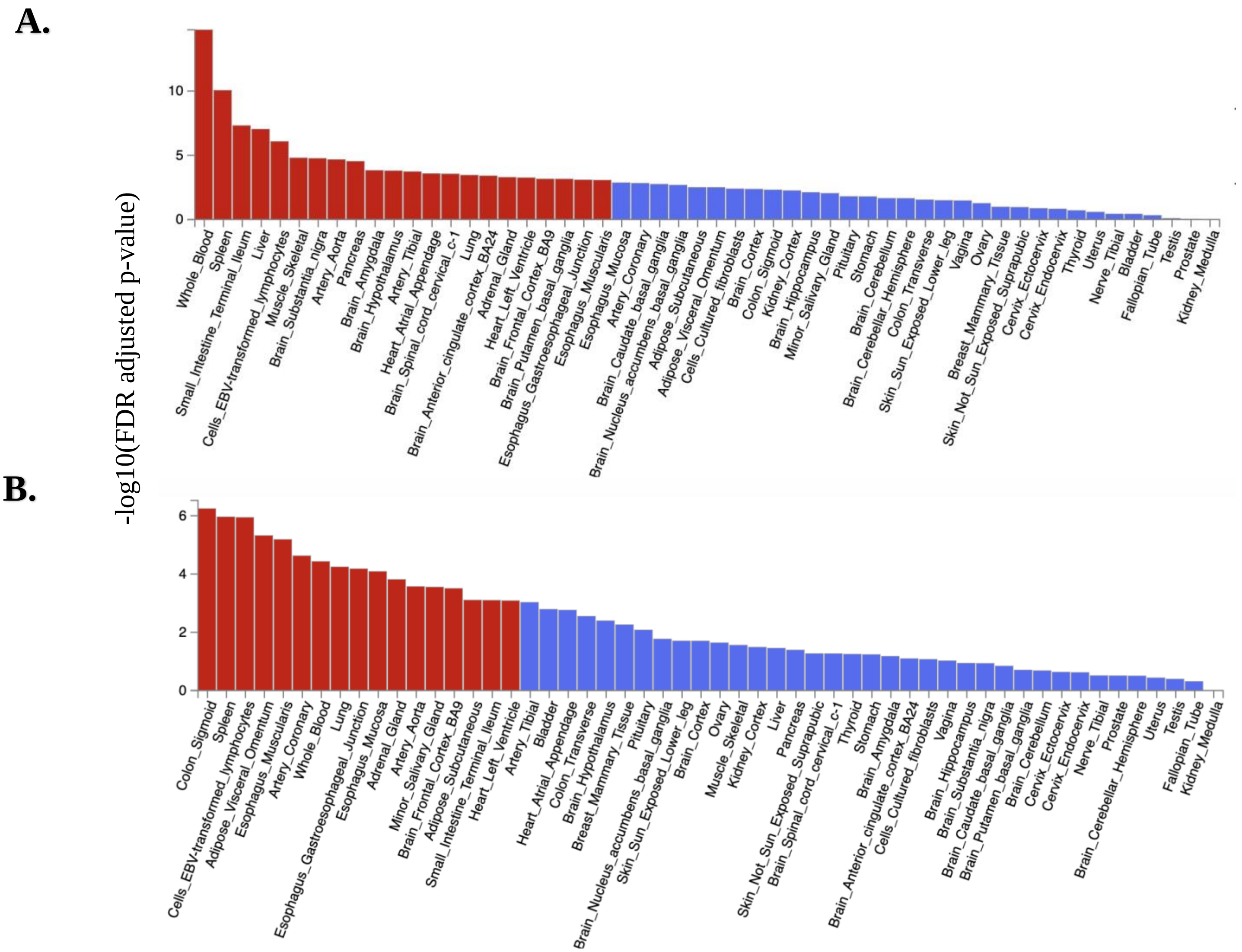


**Supplementary Figure 8.** Differential expression enrichment analysis for the selected genes in (A) gene-component 1 and (B) gene-component 2 for enrichment among pre-determined list of differentially expressed genes in each tissue of GTEx v8 and their corresponding enrichment p-value. The red bars denote the p-values for the tissues which are significant at FDR-adjusted p-value threshold of 0.001.

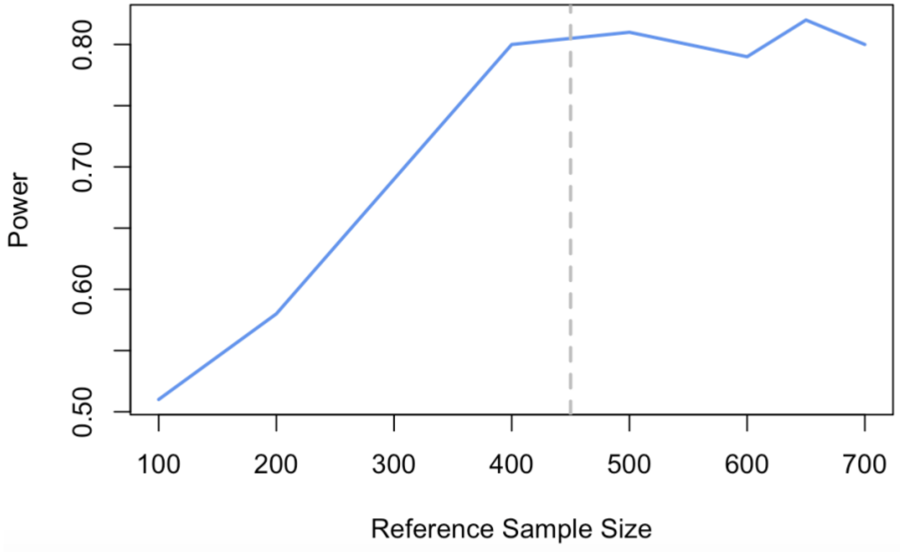


**Supplementary Figure 9. Impact of reference panel on power of ARCHIE**. The estimated empirical power of ARCHIE under sparse causal regulatory model (See Figure 2B and Simulation Model for more details), across different sample sizes of reference data to estimate coexpression matrix ($\boldsymbol{\Sigma}_{EE}$)
